# Nowcasting and Forecasting the 2022 U.S. Mpox Outbreak: Support for Public Health Decision Making and Lessons Learned

**DOI:** 10.1101/2023.04.14.23288570

**Authors:** Kelly Charniga, Zachary J. Madewell, Nina B. Masters, Jason Asher, Yoshinori Nakazawa, Ian H. Spicknall

**Author notes:** These first authors contributed equally to this article (Kelly Charniga, Zachary J. Madewell).

## Abstract

In June of 2022, the U.S. Centers for Disease Control and Prevention (CDC) Mpox Response wanted timely answers to important epidemiological questions which can now be answered more effectively through infectious disease modeling. Infectious disease models have shown to be valuable tool for decision making during outbreaks; however, model complexity often makes communicating the results and limitations of models to decision makers difficult. We performed nowcasting and forecasting for the 2022 mpox outbreak in the United States using the R package EpiNow2. We generated nowcasts/forecasts at the national level, by Census region, and for jurisdictions reporting the greatest number of mpox cases. Modeling results were shared for situational awareness within the CDC Mpox Response and publicly on the CDC website. We retrospectively evaluated forecast predictions at four key phases during the outbreak using three metrics, the weighted interval score, mean absolute error, and prediction interval coverage. We compared the performance of EpiNow2 with a naïve Bayesian generalized linear model (GLM). The EpiNow2 model had less probabilistic error than the GLM during every outbreak phase except for the early phase. We share our experiences with an existing tool for nowcasting/forecasting and highlight areas of improvement for the development of future tools. We also reflect on lessons learned regarding data quality issues and adapting modeling results for different audiences.

## Background

The 2022 mpox (formerly known as monkeypox) outbreak is the first major infectious disease outbreak since the COVID-19 pandemic and was declared a Public Health Emergency of International Concern by the World Health Organization on July 23, 2022 (1). As of April 13, 2023, a total of 86,956 confirmed cases have been reported in 110 countries and territories (2). Unlike COVID-19, mpox is a disease known to be endemic in West and Central Africa for decades; it is caused by monkeypox virus (MPXV), a zoonotic orthopoxvirus (3). Historically, classical symptoms involved fever, headache, muscle aches, fatigue, lymphadenopathy, and rash (4). Human-to-human MPXV transmission occurs through close contact with infectious material from skin lesions, respiratory secretions during prolonged face-to-face contact, and fomites, such as linens and bedding (5). The 2022 mpox outbreak began in May and spread rapidly in non-endemic countries. This outbreak was characterized by human-to-human transmission of MPXV through close physical contact (often associated with sexual activities) and has disproportionately affected gay, bisexual, and other men who have sex with men (6).

During a public health crisis such as the mpox outbreak, difficult and rapid decisions with limited available data are often required (7). Infectious disease models may assist with informing policy and practice by predicting the magnitude and duration of an outbreak or epidemic, evaluating characteristics of pathogen transmission such as transmissibility, and designing vaccination strategies, among others (8, 9). However, infectious disease models are often complex, integrating data from heterogenous sources with many parameter assumptions that are subject to uncertainty. These aspects make it challenging to effectively implement such models and communicate the results and potential limitations to decision makers, other public health partners, and the general public (10, 11).

During the COVID-19 pandemic, the state of the art of outbreak analysis advanced considerably (12). Methods and tools for estimating key epidemiological parameters, such as the effective reproduction number, *R*_*t*_, were developed and shared in real-time (13). Monitoring *R*_*t*_, the average number of secondary cases caused by a single infected individual in a large population, during an outbreak is useful for assessing transmission dynamics and evaluating the effectiveness of public health measures (e.g., vaccination, contact tracing, isolation, and quarantine) (14). Nowcasts and forecasts have been produced by numerous research groups around the globe (15-18), the results of which were instrumental for decision makers weighing possible control measures (19) such as social distancing measures. Outbreak forecasting predicts specific outcomes (e.g., number of cases, deaths, or hospitalizations) at some specific future times (e.g., weeks, months, etc.), whereas nowcasting estimates those outcomes for the current time, accounting for delays in reporting.

In this manuscript, we share our experience nowcasting and forecasting the mpox outbreak, including adapting the modeling output to different audiences. We also describe challenges faced vis-a-vis data quality, parameter estimation, and model application and propose ways to improve nowcasting and short-term forecasting efforts for future outbreaks.

## Methods

### Nowcasting/forecasting the mpox outbreak

We used data on probable and confirmed mpox cases in the United States (see “Case definition” in Supplementary methods) reported to CDC by state and local public health jurisdictions from May 17, 2022, through March 16, 2023. Data were submitted in several different formats throughout the outbreak period. These formats included: a CDC-operated call center through the Emergency Operations Center (EOC), a long and a short case report form (CRF), and the National Notifiable Diseases Surveillance System (NNDSS). Cases could have data submitted via more than one format and jurisdictions could update data on cases after initial submission (Supplementary methods). All reported data were processed in CDC’s Data Collation and Integration for Public Health Event Response (DCIPHER) platform, an instance of Palantir Foundry (Palantir Technologies Inc, Denver, CO). DCIPHER is a secure, cloud-based data integration, analytics, and situational awareness platform used by the Centers for Disease Control and Prevention (CDC), federal partners, and state, tribal, local, and territorial public health jurisdictions to collect, collaborate on, and share public health data (20). DCIPHER collates data of differing origin, structure, and purpose to provide near real-time insights into public health problems, with the goal of providing a complete picture of situational awareness.

We considered three approaches for estimating *R*_*t*_ which are implemented in the R packages EpiEstim (version 2.2-4) (21), earlyR (version 0.0.5) (22), and EpiNow2 (version 1.3.2) (23) (Supplementary methods). Initially, we used all three methods to estimate *R*_*t*_ at the national level as well as for jurisdictions reporting the highest incidence of mpox. Although estimates of the historical range of the serial interval of mpox were available at the start of the outbreak, they were based on data from the Democratic Republic of Congo, which reflected largely non-sexual household spread (24). We considered these historical parameter estimates as a starting point for early outbreak analysis, using them (along with sensitivity analyses) until new estimates were generated. Updated estimates characterized by the mean and standard deviation were needed for the global outbreak given the novel mode of transmission. In June 2022, we were able to use an estimated mean serial interval (i.e., the period of time between symptom onset in the primary case and symptom onset in the secondary case) of 9.8 days (95% credible interval [CrI]: 5.9 – 21.4) from 17 case pairs reported by the United Kingdom (6). At that time, symptom onset date was available for most reported cases, and imported cases were still contributing to a high proportion of MPXV transmission. EpiEstim results were considered the most appropriate at this stage of the outbreak because this method accounts for imported vs. locally acquired cases, has a stable codebase, is widely used, and is computationally efficient (25).

In July 2022, we started exclusively using EpiNow2, which uses a similar approach as EpiEstim (a branching process model, Supplementary methods), but it better accounts for reporting delays and incorporates multiple sources of uncertainty (13); for example, it removes noise associated with weekend effects and uses random walks for temporal smoothing. Forecasting is supported internally for *R*_*t*_, number of infections, cases by date of report, and growth rate. Unlike EpiEstim, EpiNow2 does not distinguish between imported vs. locally acquired infections. EpiNow2 is the most computationally expensive approach, requiring longer model run times (Supplementary methods). The model assumes that testing procedures, surveillance effort, and reporting delays remain constant over the estimation period. To use EpiNow2, cases by date of report must be provided as well as the generation time distribution (the time between infection of a primary and secondary case), incubation period distribution (the time between infection and symptom onset in a case), and any other delay distributions (e.g., the delay between symptom onset and report date). The model estimates the number of new cases by date of report, number of cases by their date of infection, *R*_*t*_, and time-varying growth rate. Estimates over the last 16 days of the time-series are based on partial data due to the presumption of reporting delays. Input parameters and methods for adjusting for right-truncation evolved as we learned more about the outbreak.

### Communication methods

*R*_*t*_ estimates were shared internally through Situational Reports and leadership meetings and publicly through CDC’s Technical Reports (26) and CDC’s public-facing mpox website (27). The Technical Reports were co-led by the Center for Forecasting and Outbreak Analytics (CFA) and the 2022 Multi-National Mpox Outbreak Response. Estimates were generated for distribution at least once per week. The technical reports were intended for scientific audiences. The purpose of sharing these results was to improve understanding of the outbreak and inform further scientific inquiry.

### Performance assessment methods

We chose eight dates during four key outbreak phases to retrospectively evaluate our short-term (one-week-ahead) forecasts of reported mpox cases generated from EpiNow2: 1. one month into the outbreak prior to exponential growth (June 13 and June 27); 2. during exponential growth (July 5); 3. near the outbreak peak (July 27); and 4. during the declining phase (September 6, September 19, October 11, and December 5). Ideally, the same day of the week would be used, but some historical versions of the dataset were not available for this analysis and some dates fell on national holidays which may have introduced additional delays. Like the real-time analyses, we used rash onset date as the first reference date to define the reporting delay distribution for all eight time points, while the second reference date changed over time (Table S1).

We used three metrics to evaluate the forecasts. Our primary metric was the weighted interval score (WIS). For each of the eight time points considered, WIS was computed for each daily prediction and averaged across the seven-day forecast. The WIS measures the consistency of a group of prediction intervals with an observed value (probabilistic accuracy). The WIS is positive, and lower values correspond to smaller error (Supplementary methods) (15). To evaluate the error in the forecast’s point estimate, we used the mean absolute error (MAE), which was computed as 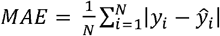, where *y*_*i*_ is the observed number of mpox cases on day *i, ŷ*_*i*_ is the median forecast on day *i*, and N = 7 (15). We also used prediction interval (PI) coverage rates, which check the degree to which the model provides calibrated predictions. Coverage rates are calculated by determining the proportion of times the 50% or 90% PIs included the observed value (for example, a well-calibrated forecast would have a 50% PI coverage close to 0.50. Also see Supplementary methods) (15).

We compared the performance of EpiNow2 with a naïve Bayesian generalized linear model (GLM, Supplementary methods). We calculated the relative WIS and relative MAE for EpiNow2 and the GLM as 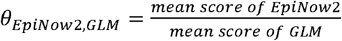, where the mean score is the average of the models’ performance (WIS or MAE) over all eight dates evaluated. If θ_*EpiNow2,GLM*_ was less than 1, that indicated the forecasts generated by EpiNow2 had less error than the GLM, whereas θ_*EpiNow2,GLM*_ > 1 indicated EpiNow2 performed worse.

For both EpiNow2 and the GLM, we removed recent cases (defined as cases reported in the last 3 – 5 days) from the time series to adjust for right truncation of the data for all four outbreak phases (Table S1). Forecasts were evaluated using mpox data as of March 16, 2023. We used the most recent version of event date as the basis for the comparison.

## Results

### Challenges of nowcasting/forecasting

It is voluntary for jurisdictions to report mpox cases to CDC, with only minimal data needed to submit a case report form (e.g., case ID and reporting jurisdiction) in part, because not all cases may be reached or fully investigated. CDC asks jurisdictions to collect and report additional data variables to achieve situational awareness and surveillance goals. The number of variables on the case report form was decreased to reduce reporting burden. Despite these efforts, jurisdictional case surveillance systems may not have aligned to CDC’s requested data variables, and jurisdictions may choose to limit what data are shared with CDC based on local reporting practices. Received data were subjected to additional manual data cleaning to standardize formats and correct obvious data entry errors. As a result, even key data variables such as demographic characteristics (e.g., age, race/ethnicity, HIV status, vaccination) were not consistently available across all jurisdictions and time periods, precluding detailed sub-analyses. For example, out of 29,921 cases in DCIPHER through December 31, 2022, 21,480 (71.8%) were missing HIV status, 16,474 (55.1%) were missing smallpox vaccination, 3,913 (13.1%) were missing gender identity, 3,172 (10.6%) were missing race, 2,928 (9.8%) were missing ethnicity, and 250 (0.8%) were missing age. The timing and frequency of data submission varied between jurisdictions and changed over the course of the outbreak. Some jurisdictions reported case data in near real-time whereas others submitted a large number of cases all at once, the latter of which caused large, artificial spikes in the time-series. There were instances of duplicate cases being reported from several jurisdictions which may be attributed to the changes in reporting processes. Spurious cases at the end of the time series had to be investigated (and usually removed) because they artificially inflated the nowcasts/forecasts. These data issues required us to monitor the model output closely and modify the code as needed.

In early July 2022, reporting of mpox cases to CDC started to lag in some jurisdictions, especially those most affected by the outbreak. These few jurisdictions were publicly reporting more cases on their websites than what CDC had received reports for. This led to a lengthy case reconciliation process during which case data uncertainty prevented it from being used for nowcasting/forecasting at the national level. Also in July, an increasing proportion of cases were reported with missing symptom onset dates (from 26% to 53% for rash onset date between June 13 and July 5). To ensure each case had a date associated with it for plotting epidemic curves, a new event date field was created which we started using for nowcasting/forecasting. The new calculated date field selected the best available date among possible date fields based on the following priority, ordered from most to least preferable: orthopoxvirus test date, date of call to the call center, date the short CRF was created, and the long CRF timestamp (Figure S1). The date in which the record was created was least preferred due to artificial spikes in the time series caused by bulk uploading data. In September 2022, a different date was adopted by the response for reporting case data. This date field was defined as the earliest among all available dates for a case including symptom onset, which facilitated improved visualization of epidemic curves. However, this new date presented challenges for its use in the EpiNow2 framework because the delay from symptom onset to this new date would have more variation than the delay using the original event date, including a delay of zero for some cases. Thus, we worked to create a new event date field specifically for nowcasting/forecasting which was similar to the original event date. The definition was expanded to include dates available in NNDSS data.

### Successes of nowcasting/forecasting

During the case reconciliation process in July, publicly available data through health department websites was used for subnational analyses [e.g., California (28) and New York City (29). We used WebPlotDigitizer (30) to extract time series data from pdfs when the underlying data were not available for download.

In October, we updated estimates of the serial interval for rash onset of 7.0 days (95% CrI 5.8 – 8.4) from 40 case pairs and incubation period for rash onset of 7.5 days (95% CrI 6.0 – 9.8) from 35 U.S. case-patients and used those as model inputs for EpiNow2 (31). These data were obtained through the collaboration of several U.S. jurisdictions on a special study. The estimated serial interval for the 2022 outbreak was on the lower end of the historical range observed in the Democratic Republic of Congo (7 – 23 days) (24).

### Adapting model output and communicating nowcasts/forecasts

We adapted the presentation of our results for a scientific/technical audience and the general public. The default plots from EpiNow2 included three panels: cases by date of report, cases by date of infection, and *R*_*t*_. Green represented estimates based on complete data, orange represented estimates based on partial data, and purple represented the forecast (the default is seven days). Gray bars in the top panel showed the actual time series of reported cases, while gray bars in the middle panel showed the back-calculated infection time series. For the Technical Reports, Situational Reports, and response updates meetings, we removed the middle panel (Figure 1) (26). For the website, we only showed *R*_*t*_, removed the forecast, and removed the 20% credible intervals to minimize confusion (Figure 2) (27). We included a simple description of the plot that could be understood by non-experts. In accordance with CDC’s Data Modernization Initiative, a national effort aimed at modernizing state and national core data and surveillance infrastructure (32), data for the underlying plots were made available for download as comma-separated values (csv) files with the Technical Reports.

**Figure 1.**
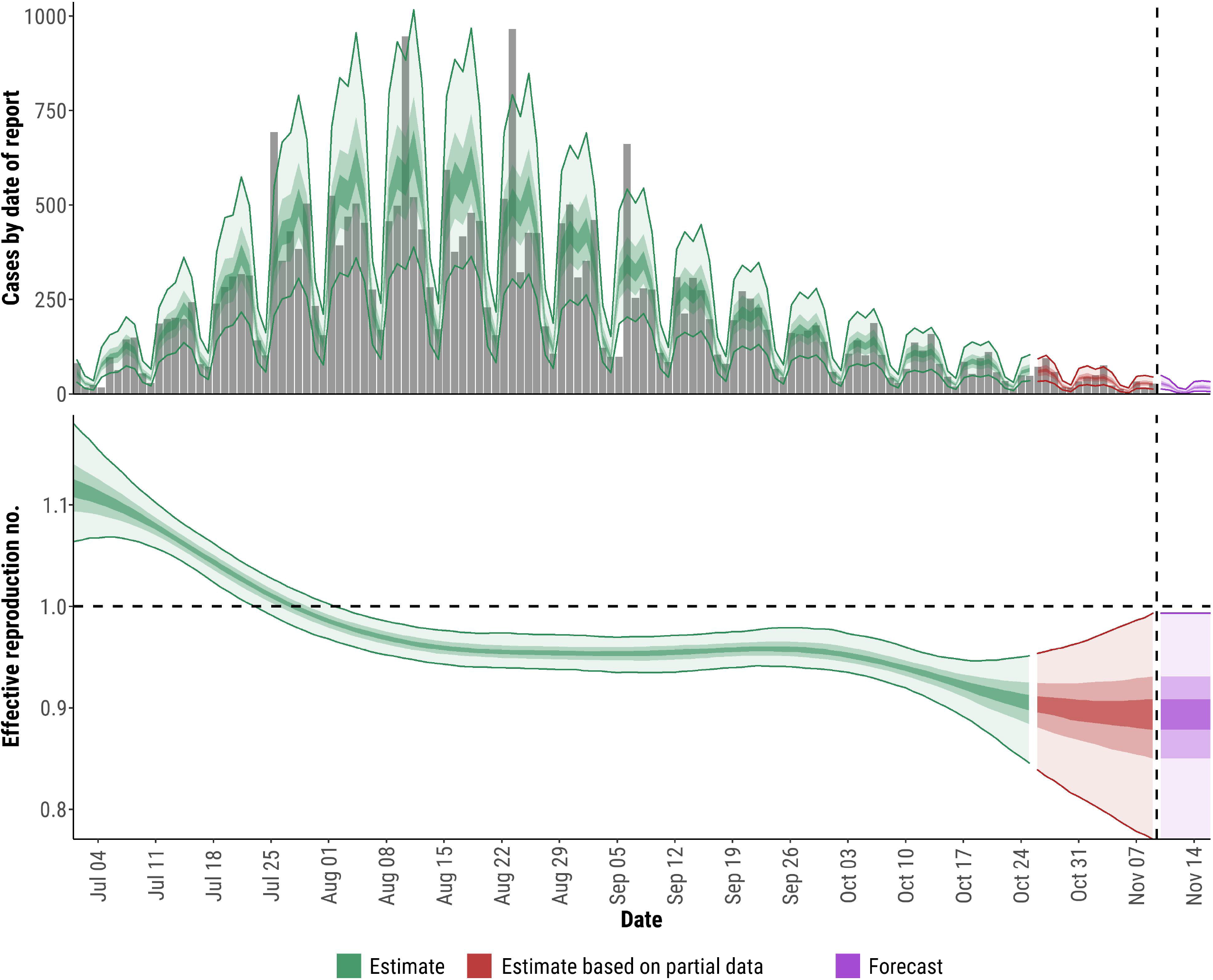
Effective reproduction number estimates for the U.S. 2022 mpox outbreak intended for a technical/scientific audience. The top panel shows estimates of cases by date of report with actual cases shown by gray bars. The bottom panel shows estimates of the effective reproduction number by date. In all panels, shaded regions reflect 90%, 50%, and 20% credible intervals in order from lightest to darkest. Green shows estimates, red shows estimates based on partial data, and purple shows forecasts. Event date is determined by a hierarchy across the different data streams where priority is given to diagnosis date, orthopoxvirus test date, orthopoxvirus test confirmation date, case investigation start date, orthopoxvirus sample collection date, date of call to CDC call center, report date (to public health department, county, or state), date CDC announced case, and the date the case was entered into DCIPHER, in that order.

**Figure 2.**
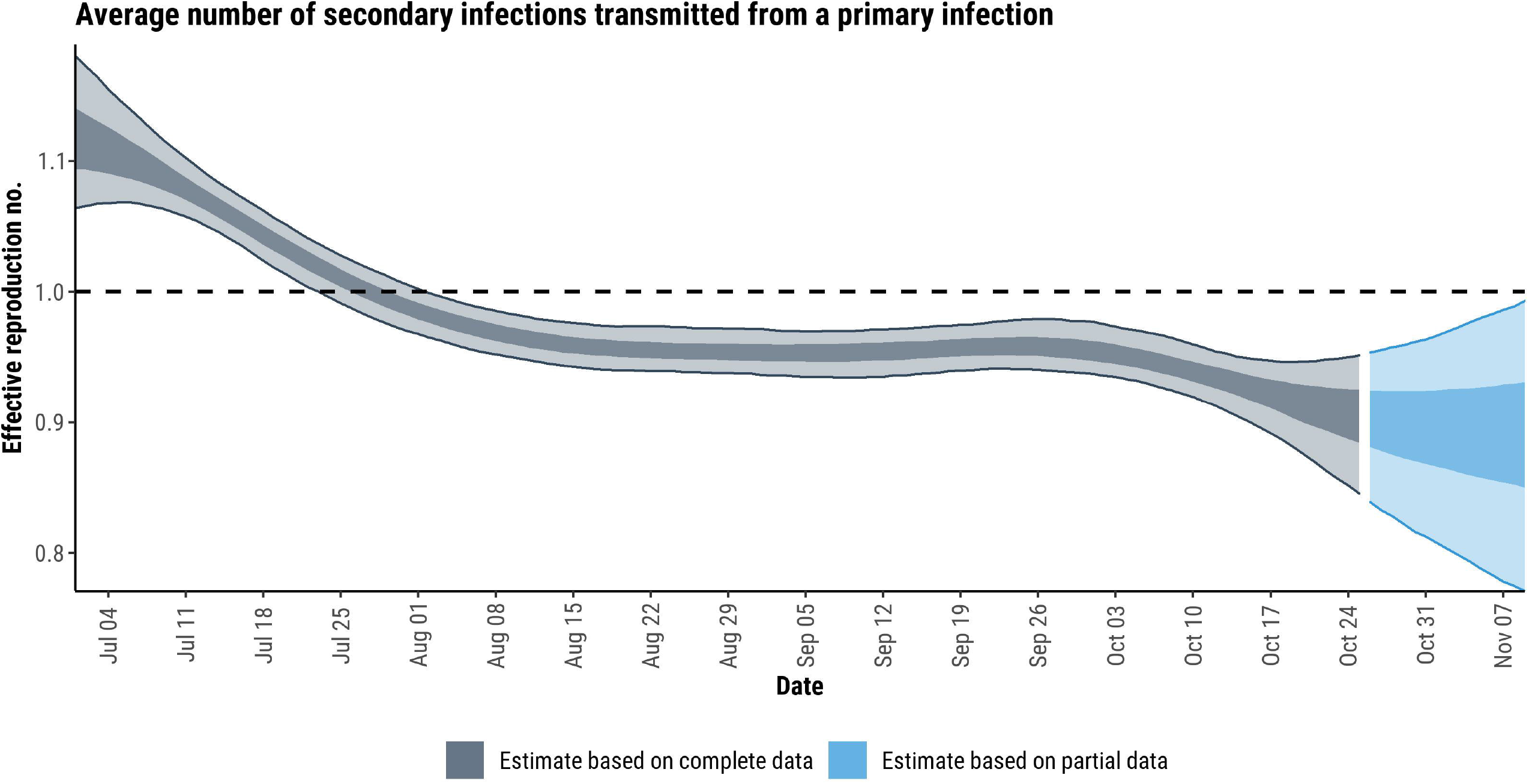
Effective reproduction number (*R*_*t*_) estimates for the U.S. 2022 mpox outbreak intended for the general public. The graph shows the *R*_*t*_ estimation over time based on complete data (gray) or partial data (blue). The most recent data are considered incomplete due to delays in reporting mpox cases. As a result, there is more uncertainty associated with the most recent *R*_*t*_ estimates. *R*_*t*_ > 1 means the epidemic is growing. *R*_*t*_ < 1 means the epidemic is shrinking. Shading represents the 50% and 90% credible intervals (uncertainty in the estimates)

Sub-national analyses revealed some differences between regions regarding the start of the outbreak, when it peaked, and how long it lasted (Figures 3 – 4). For example, Figure 4 demonstrates a later introduction date and slightly longer tail for Texas compared to other jurisdictions.

**Figure 3.**
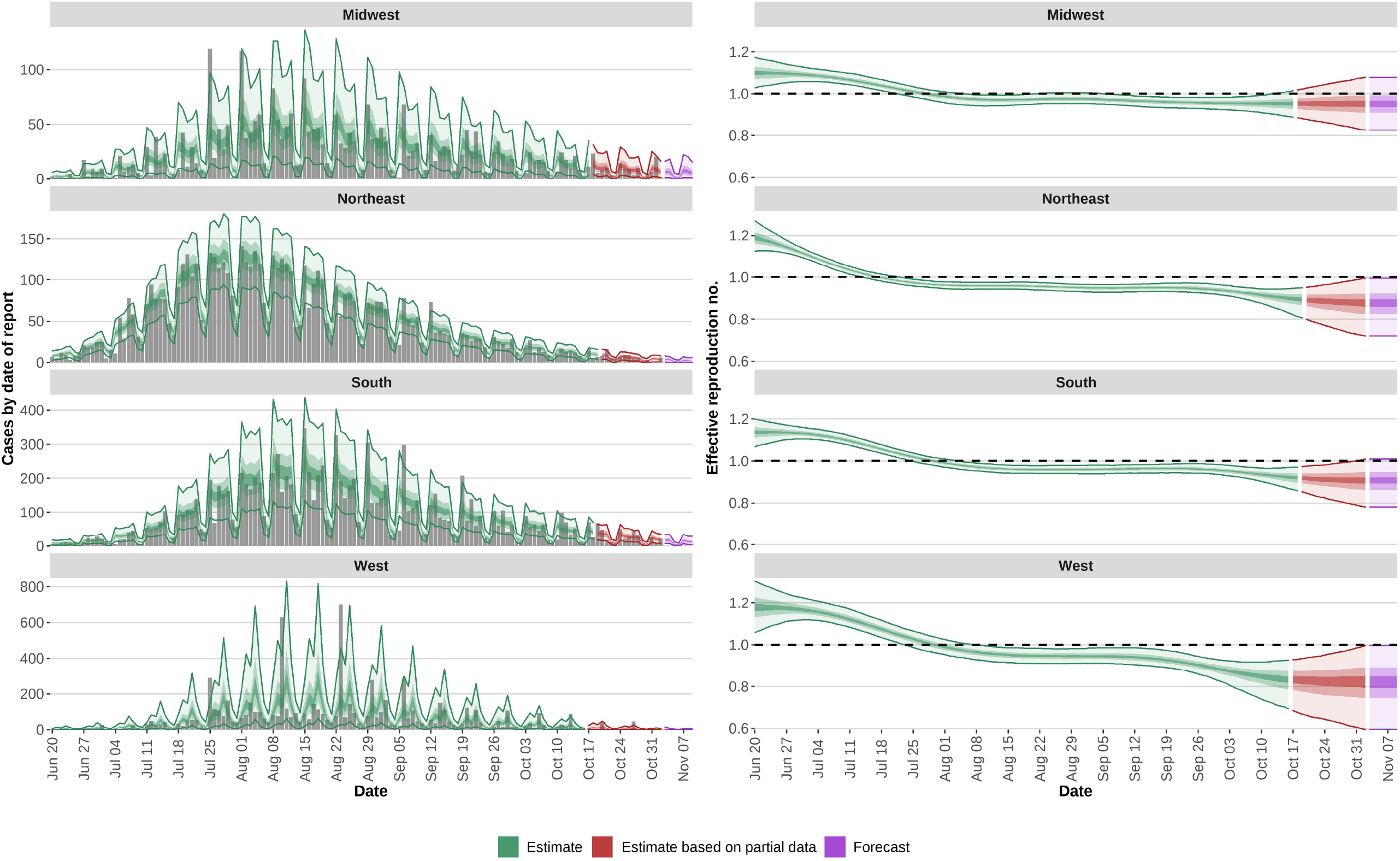
Effective reproduction number estimates for the 2022 mpox outbreak in four U.S. Census regions. The left panels show estimates of cases by date of report with actual cases shown by gray bars. The right panels show estimates of the effective reproduction number by date. In all panels, shaded regions reflect 90%, 50%, and 20% credible intervals in order from lightest to darkest. Green shows estimates, red shows estimates based on partial data, and purple shows forecasts. Event date is determined by a hierarchy across the different data streams where priority is given to diagnosis date, orthopoxvirus test date, orthopoxvirus test confirmation date, case investigation start date, orthopoxvirus sample collection date, date of call to CDC call center, report date (to public health department, county, or state), date CDC announced case, and the date the case was entered into DCIPHER, in that order.

**Figure 4.**
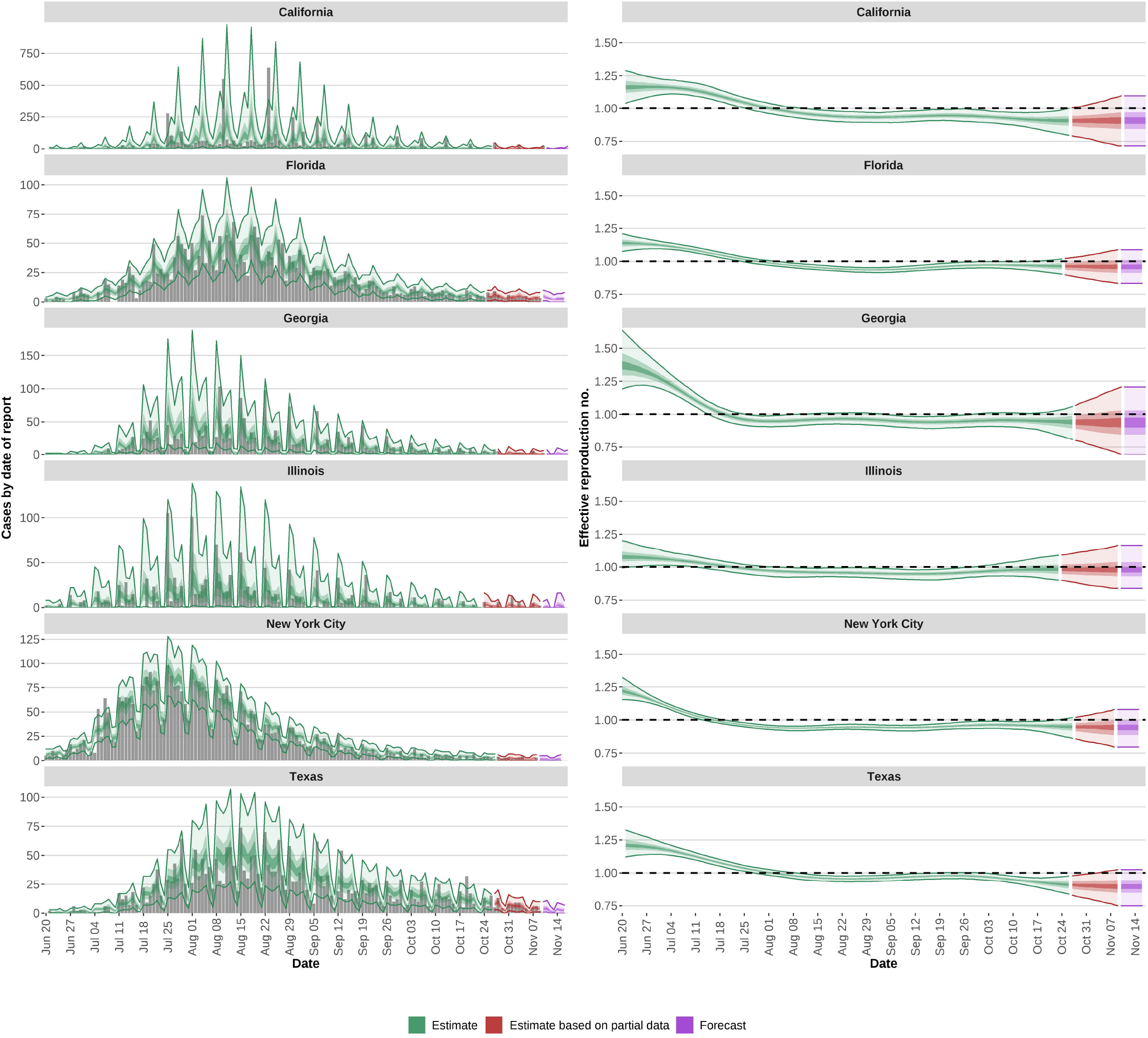
Effective reproduction number estimates of the 2022 mpox outbreak for the six jurisdictions in the U.S. with the highest case counts. The left panels show estimates of cases by date of report with actual cases shown by gray bars. The right panels show estimates of the effective reproduction number by date. In all panels, shaded regions reflect 90%, 50%, and 20% credible intervals in order from lightest to darkest. Green shows estimates, red shows estimates based on partial data, and purple shows forecasts. Event date is determined by a hierarchy across the different data streams where priority is given to diagnosis date, orthopoxvirus test date, orthopoxvirus test confirmation date, case investigation start date, orthopoxvirus sample collection date, date of call to CDC call center, report date (to public health department, county, or state), date CDC announced case, and the date the case was entered into DCIPHER, in that order.

### Performance assessment

The GLM had lower WIS compared to EpiNow2 for early phase of the outbreak (Table 1); however, during all other phases, EpiNow2 had a slight advantage. The relative WIS was 0.89 over all eight time points considered, indicating that EpiNow2 had on average 11% less probabilistic error than the GLM.

**Table 1.** Evaluation of short-term (one-week-ahead) forecasts of reported mpox cases during the 2022 U.S. outbreak. For WIS, bold indicates where one model performed better than the other.

| Forecast date | Outbreak phase | EpiNow2 |  |  |  | Bayesian GLM with negative binomial |  |  |  |
| --- | --- | --- | --- | --- | --- | --- | --- | --- | --- |
|  |  | 90% PI coverage | 50% PI coverage | WIS | MAE | 90% PI coverage | 50% PI coverage | WIS | MAE |
| Monday, June 13 | Early | 1 | 0.71 | 33.8 | 4.3 | 0.86 | 0.57 | **6.4** | 5.1 |
| Monday, June 27 | Early | 0.86 | 0 | 34.0 | 29.0 | 0.86 | 0.14 | **24.7** | 26.9 |
| Tuesday, July 5 | Exponential growth | 0.86 | 0.71 | **35.6** | 32.4 | 1 | 0.57 | 38.5 | 34.1 |
| Wednesday, July 27 | Peak | 1 | 0.57 | **137.0** | 107.9 | 1 | 0.57 | 184.3 | 94.4 |
| Tuesday, September 6 | Decline | 0.86 | 0.71 | **107.6** | 84.3 | 0.86 | 0.43 | 117.1 | 86.8 |
| Monday, September 19 | Decline | 0.86 | 0.43 | **61.2** | 52.4 | 0.71 | 0.43 | 81.8 | 65.9 |
| Tuesday, October 11 | Decline | 0.43 | 0.29 | **30.6** | 33.4 | 0.57 | 0.14 | 38.3 | 41.9 |
| Monday, December 5 | Decline | 1 | 0.71 | **4.3** | 3.1 | 1 | 0.71 | 6.4 | 4.6 |

359 PI: prediction interval; WIS: weighted interval score; MAE: mean absolute error; GLM: generalized linear model

EpiNow2 had lower MAE than the GLM for six out of eight time points, but performance was similar: the relative MAE was 0.96. In other words, EpiNow2 had only 4% less point error than the GLM.

Overall, predictions were moderately well calibrated. For the 90% PI, EpiNow2 achieved coverage rates within 10% of the desired coverage level for seven out of eight time points compared to six out of eight for the GLM. For the 50% PI, EpiNow2 achieved coverage rates within 10% of the desired coverage level for only two out of eight time points compared to five out of eight for the GLM.

Qualitative results of the nowcasts/forecasts are shown in Figure S2. The 90% CrIs from EpiNow2 were very wide for the early phase of the outbreak, while the GLM had large uncertainty around the outbreak’s peak. Both models underestimated reported mpox cases for the seventh time point on October 11 which could be due to a discrepancy in the data available at the time versus the ground-truth data (Figure S3). This time point had the lowest PI coverage rates.

## Discussion

We performed nowcasting/forecasting to inform the U.S. response to the 2022 mpox outbreak in real-time. Validation showed that the method implemented in EpiNow2 predicted case counts reasonably well, but improvements are needed around key time periods such as the outbreak’s peak. One reason that the nowcasts/forecasts did not always align with reality is because the definition of event date changed over time, while the study data were constructed the most recent version of the event date field. We found a higher WIS for EpiNow2 in the early phase of the outbreak compared to the GLM which could be due to choices of priors for parameters (e.g., wide intervals for *R*_*t*_).

Subnational analyses allowed us to better understand the spatial heterogeneity of the epidemic which may be attributed to differences between jurisdictions in terms of composition (e.g., population age structure, density, and contact patterns) and public health activities (e.g., vaccination, surveillance methods, frequency of testing) (33) as well as the timing and frequency of case reporting. One limitation of nowcasting/forecasting at the subregional or jurisdictional level is that the effects of bulk uploads are more apparent, resulting in greater uncertainty (wider credible intervals). Another limitation is that movement between jurisdictions could have a greater impact on subnational estimates, as mobility is not accounted for in our approach. Finally, some jurisdictions stopped reporting rash onset date, which decreased the sample size available for estimating the reporting delay distribution over time.

### Data Quality

Nowcasting/forecasting methods perform best when the underlying surveillance data are accurate, timely, and complete, but they are often sub-optimal and variable as the outbreak evolves; while data may improve as an outbreak progresses, they may re-deteriorate once the outbreak slows and intensity of effort is low. Fortunately, the quality and frequency of data improved over the course of the U.S. mpox outbreak. Communicating with specific jurisdictions about our priority dates for modeling improved data quality. These prompts to the jurisdictions need to be continued regularly throughout the outbreak. Close collaboration between epidemiologists/modelers and informaticians, including the use of an issue tracking system in DCIPHER, also facilitated quick investigation and resolution of data errors.

### EpiNow2 Limitations

The main limitation of EpiNow2 is its steep learning curve due to limited documentation of package functions and few reports of its application to other outbreaks. Increasing commenting in the code, creating more tutorials or vignettes, and developing a graphical user interface could help.

Another limitation is the long computing time required for the analyses. We were able to increase computational efficiency by running the model on multiple cores in parallel, but the processing time became particularly cumbersome if an analysis needed to be repeated. In the future, cloud-based computing could be used to obtain more consistent and faster model run times.

There were also instances of unusually long run times whereby the first two Markov chain Monte Carlo (MCMC) chains performed as expected, but subsequent chains never finished processing. Some MCMC convergence issues were resolved by reducing the parameter fitting period (e.g., truncating the beginning of the time series). One study reported that EpiNow2 estimates are more reliable when case numbers at each time step are large and there are at least 14 timepoints without zeroes (34). Large daily fluctuations and limited case counts could substantially affect model estimates, which should be interpreted with caution.

Another limitation is that the method we used does not account for under-ascertainment, which occurs when not all infections are diagnosed and reported as cases of the disease to the surveillance system. The under-ascertainment rate is needed to understand the true burden of disease caused by the outbreak; however, current estimates for the U.S. mpox outbreak are lacking. Indirect evidence from a recent modeling study (35) suggests that 65% of mpox infections were diagnosed and reported in Washington, D.C. However, the model was not designed to measure the under-ascertainment rate (Patrick Clay, personal communication, March 10, 2023), and this quantity should be assessed by other methods (e.g., models specifically designed to assess under-ascertainment, serological surveys, and community-based surveys).

### Strategies for Forecasting the Next Outbreak

For the next outbreak, it is important for CDC to develop strategies for regularly capturing and storing snapshots of surveillance data which remain easily accessible for systematic analysis. For routine case-based surveillance of notifiable diseases, such as rabies, most analyses are performed only after the data have undergone a rigorous and routine reconciliation and closeout process by data submitters with further validation by CDC surveillance epidemiologists; however, timely outbreak response decision support does not allow for such processes. Instead, jurisdictions are asked to submit available case data in near real-time and submit additional data or corrections to data entry errors as time and resources permit. Snapshots of the surveillance data were saved in an ad hoc manner (by exporting data on a particular day and saving a csv file locally), and as a consequence, a complete history of the data is not available, especially around key points in the outbreak, such as the peak. A complete history would help to understand key delay distributions and other quirks (e.g., backfilling and revision of reference dates) involved in the data-generating process. Understanding the data generating process is crucial for the improvement of methods and tools for nowcasting/forecasting and aligns with one of the five priorities of CDC’s Data Modernization Initiative (Accelerating Data for Action: Tapping into more data sources, promoting health equity, and increasing capacities for scalable outbreak response, forecasting, and predictive analytics) (32). In the future, the process of saving snapshots of the data could be automated.

Ensemble models have been used for a variety of infectious disease outbreaks, such as COVID-19 (15, 36), Zika (37), influenza (38), and Ebola (39). Ensembles combine predictions from several models that use different methodology and sometimes input data. Because some models overpredict, while others underpredict, ensemble models often outperform individual models over time. In the future, we may consider using at least two simpler models and comparing them.

One potentially useful addition to EpiNow2 and other currently available tools for nowcasting/forecasting outbreaks would be flexibility in handling dates. We frequently encountered missing dates for cases in the mpox surveillance data. Ideally, a method or tool would be able to keep track of multiple dates for a case and estimate missing dates based on the full distribution of dates across all cases. Epinowcast is a new hierarchical nowcasting package that enables more flexibility in adjusting for truncated data (40). Novel nowcasting approaches use hierarchical generalized additive models, which can provide even more flexibility to modify the model in real-time to the evolving data environment (41). Another improvement would be to reduce the time required to run the analyses. Rather than focusing on the efficiency of the MCMC algorithm, computation time could be reduced if the model only needed to be run on the new data. Finally, the imputed time series of cases by symptom onset date would be a useful data visualization output that is not currently available in EpiNow2. As described above, defining the date field for the presentation of epidemic curves was a challenge in the mpox outbreak and having an imputed symptom onset date for each case would have been useful for comparison purposes.

CFA played an important advisory role in our nowcasting/forecasting efforts. CFA produces models and forecasts to characterize the state of an outbreak and its course, inform public health decision makers on potential consequences of deploying control measures, and support innovation to continuously improve the science of outbreak analytics and modeling (42). In the future, CFA plans to create new tools for outbreak analysis and modeling. CFA could also serve as a link between CDC modelers and jurisdictions with modeling capacity to share experiences and code. Technical Reports represent a new way for CDC to share timely information with the federal government, state and local leaders, and scientists in academia and industry. Technical Reports have been well received within and outside CDC (43-45) and their publication aligns with CDC’s current restructuring efforts aimed at making the agency more response ready, including sharing science and data faster (46).

## Conclusion

Real-time estimation of *R*_*t*_ as well as nowcasting/forecasting is one method for determining the extent to which current public health measures are effective and/or need to be modified but is subject to limitations. The quality and timeliness of reported data pose challenges to these analyses. Ease of use, model computing time, and ability to handle multiple dates are priorities for consideration in the development of future nowcasting/forecasting tools. A naïve model may be superior to a complex one, such as EpiNow2, during the early phase of an outbreak when data scarcity causes *R*_*t*_ to be largely unconstrained, especially once reporting delays are considered. Future outbreak response activities could be enhanced through inclusion of clear and consistent communication about modeling outputs as well as close collaboration between modeling and informatics/data teams.

## Data Availability

Data and code to run the nowcasts/forecasts and perform model validation will be available on GitHub following publication in a peer-reviewed journal.

## Acknowledgements

We thank all public health professionals involved in reporting mpox cases to CDC. We also acknowledge the Mpox Response Data Analytics and Visualization Task Force Informatics Team for data management and support, and we thank Dr. Sam Abbott for helpful discussions about EpiNow2 methods.

## Disclaimer

The findings and conclusions in this report are those of the authors and do not necessarily represent the official position of the Centers for Disease Control and Prevention, U.S. Department of Health and Human Services.

## Ethics statement

This activity was reviewed by CDC and was conducted consistent with applicable federal law and CDC policy (45 C.F.R. part 46, 21 C.F.R. part 56; 42 U.S.C. Sect. 241(d); 5 U.S.C. Sect. 552a; 44 U.S.C. Sect. 3501 et seq).

## Author contributions

Conceptualization (KC, YN), data curation (KC, JA), formal analysis (KC, ZJM), investigation (KC, ZJM), methodology (KC, ZJM, JA), project administration (KC), software (KC, ZJM, NBM), supervision (JA, YN, IHS), validation (KC, ZJM), visualization (KC, ZJM), writing – original draft preparation (KC, ZJM), writing – review & editing (KC, ZJM, NBM, JA, YN, IHS).

