## Appendix for "Nowcasting and Forecasting the 2022 U.S. Mpox Outbreak: Support for Public Health Decision Making and Lessons Learned"

**Supplementary methods**

**Supplementary tables**

**Table S1.** Input parameters and truncation adjustment for nowcasting/forecasting by outbreak phase.

**Supplementary figures**

**Figure S1.** Hierarchy of event dates used for nowcasting/forecasting the 2022 U.S. mpox outbreak.

**Figure S2.** Retrospective evaluation of nowcasts/forecasts of mpox cases in the United States.

**Figure S3.** Time series of reported mpox cases in the United States used for nowcasting/forecasting.

**References**

**Disclaimer:** The findings and conclusions in this report are those of the authors and do not necessarily represent the official position of the Centers for Disease Control and Prevention, U.S. Department of Health and Human Services.

**Supplementary methods**

**Case definitions.** A probable case of mpox was defined as having no suspicion of other recent orthopoxviru*s* exposure AND demonstration of orthopoxvirus DNA by polymerase chain reaction of a clinical specimen OR orthopoxvirus using immuno or electron microscopy testing methods OR demonstration of detectable levels of anti-orthopoxvirus IgM antibody during the period of 4 to 56 days after rash onset (1). A confirmed case of mpox was defined as demonstration of the presence of MPXV DNA by polymerase chain reaction testing or Next-Generation sequencing of a clinical specimen OR isolation of MPXV in culture from a clinical specimen (1).

**Data sources.** The Centers for Disease Control and Prevention (CDC) call center was set up on May 19 to support jurisdictions in evaluating possible cases of mpox, including recommendations for clinical diagnosis and orthopoxvirus testing as well as linking cases across jurisdictions (2). The call center was phased out in late July 2022. The long CRF was implemented on June 2, 2022, consisting of 223 questions with skip logic. Data were collected on domestic mpox cases to understand the epidemiology, clinical presentation, and severity of disease as well as inform public health interventions, identify risk factors for infection, and interrupt disease transmission. The short CRF became available on July 1, 2022. It consisted of a subset of questions from the original (long) CRF. The National Notifiable Diseases Surveillance System (NNDSS) is the national system used to report approximately 120 notifiable diseases and conditions to CDC (3). It was approved as a mechanism for reporting mpox cases in August 2022. CDC used a phased approach for onboarding jurisdictions to NNDSS. As of early November, eight states had been onboarded.

**Alternative tools for nowcasting/forecasting.** EpiEstim relies on a branching process model. It was first developed in 2013 and is widely used for outbreak analysis (4, 5). *R_t_* is estimated as the ratio of the number of new infections produced at time *t*, *I_t_*, to the total infectiousness of infected people at time *t*: $\sum_{s=1}^{t} I_{t-s}w_{s}$, where *w_s_* is the probability mass function of the generation time. Estimation is performed over user-defined time windows, and the length of the time window determines the smoothness of the estimates. EpiEstim can distinguish between imported and locally transmitted cases and can be used in conjunction with the projections R package(version 0.5.4) (6) to forecast future epidemic trajectories. To use the package, the user must provide data on daily incidence by date of symptom onset and specify the serial interval.

EarlyR is a simplified version of EpiEstim that implements a likelihood-based estimation of *R_t_* using a branching process with a Poisson likelihood. It is better suited to estimate *R_t_* at the beginning of an outbreak when few cases have been reported. Epidemic trajectories can also be forecasted using the projections R package (6). To use earlyR, the user must provide data on daily incidence by date of symptom onset and specify the serial interval.

**Adjusting for right truncation.** Adjusting for right truncation (the absence of recent infections in the data) is considered best practice for real-time outbreak analysis. If right truncation is not adjusted for, nowcasts/forecasts will seem more optimistic because recent infections will not have been reported yet (7). The estimate_truncation() function in EpiNow2 estimates a truncation distribution from “snapshots” of the data over time (8). A snapshot is a version of the dataset that was available at a previous time point. The output from this function can be passed as an argument to the epinow() function to adjust for right truncation. We used this approach initially with EpiNow2. When reporting started to stabilize, we changed our approach: instead of using estimate_truncation(), we removed cases from the time series that were first entered into the DCIPHER platform during the previous 3 – 5 days. This method reduced computational time and avoided data errors present in the snapshots that were later corrected.

When retrospectively evaluating forecasts, we did not always have data available for Monday, so we ran the data using the previous Friday’s data for consistency.

**Final EpiNow2 methods.** The methods in this section correspond to those used near the end of the outbreak (from September 28, 2022). They were used to produce Figures 1 – 4 in the main text.

We used the EpiNow2 package to estimate *R_t_* during the mpox outbreak in the United States. We included all probable and confirmed mpox cases. We evaluated *R_t_* separately for the United States, four Census regions (9), and seven jurisdictions. As mentioned in the main text, EpiNow2 requires three distributions: 1. the generation time, 2. the incubation period, and 3. any other delays. We used event date as the date field for our time series (Figure S1, right panel). We approximated the generation time using an estimate of the serial interval, or time between symptom onset in a primary case and symptom onset in the secondary case. For this analysis, we used the mean serial interval for rash onset of 7.0 days (95% credible interval [CrI]: 5.8 – 8.4) and standard deviation of 4.2 (95% CrI: 3.2 – 5.6) from 40 primary/secondary case pairs across 12 U.S. jurisdictions (10). We also used the mean incubation period from exposure to rash onset for 35 cases of 7.5 days (95% CrI: 6.0 – 9.8) and standard deviation 4.9 (95% CrI: 3.2 – 8.8) (10). The delay from rash onset to reporting was estimated by fitting a bootstrapped log-normal distribution with a maximum delay of 29 days for a subset of cases with both dates available.

The model was fitted to the time-series of incident reported cases per day using Markov chain Monte Carlo (MCMC) methods, implemented in Stan (11). We ran three chains and drew 2,000 samples for each chain. We also generated short-term estimates of future incidence using the default horizon of seven days. We reported the estimated 20%, 50%, and 90% CrIs of the posterior distributions for all output parameters.

To adjust for right-truncation, we removed cases reporting during the most recent 3 – 5 days from the time series.

We used R version 4.1.1 for all analyses.

**Computational needs.** We initially ran EpiNow2 locally on our laptops, pulling data from DCIPHER into R using an application programming interface (API). Our laptops use Windows operating system and have four cores. We ran three MCMC chains in parallel with 2,000 samples each. Run times ranged from approximately 10 – 40 minutes, depending on how many other tasks we were using our laptops for at the same time (e.g., video calls).

We were encouraged to run the nowcasts/forecasts in DCIPHER so that the output could be more easily shared across the other teams involved in the response. DCIPHER can run R code through SparkR. It has features such as scheduling builds, which run code automatically at designated times, and in theory, faster model run times because the code would be running on an external server. In practice, we encountered several difficulties after transferring our code to a DCIPHER workbook. The primary issue was that our model builds suffered from long queuing times in the code workbook, despite no other code being run at the same time. This problem was never solved. In our experience, debugging code in DCIPHER was considerably more difficult than in R; it took more time, and the error messages were often uninformative. Ultimately, we went back to running models locally in R. DCIPHER could be used for nowcasting/forecasting future outbreaks, but ideally the problems we experienced would need to be addressed first.

**Calculating weighted interval score and prediction interval coverage.** For quantiles of a forecast distribution, *F*, an observation, *y*, and uncertainty level, *α*, a single interval score is defined as

$${IS}_{\alpha}\left( F,y \right)=\left( u-l \right)+\frac{2}{\alpha}\cdot\left( l-y \right)\cdot1\left( y<l \right)+\frac{2}{\alpha}\cdot\left( y-u \right)\cdot1\left( y>u \right)$$

where 1($\cdot$) is the indicator function and $l$ and $u$ are the $\frac{\alpha}{2}$ and $1-\frac{\alpha}{2}$ quantiles of *F*(12). This equation can be broken down into three main parts: dispersion, underprediction, and overprediction, in that order. To summarize accuracy across a predictive distribution, we can calculate a weighted sum of interval scores. The weighted interval score (WIS) is therefore defined as

$${WIS}_{\alpha_{0:K}}\left( F,y \right)=\frac{1}{K+1/2}\cdot\left( w_{0}\cdot\left| y-m \right|+\sum_{k=1}^{K} w_{k}\cdot{IS}_{\alpha_{k}}\left( F,y \right) \right)$$

where $w_{k}=\frac{\alpha_{k}}{2}$ for $k=1\ldots, K$, $w_{0}=1/2$, and $m$ is the median of the distribution. We used $K=3$ interval scores ($\alpha=0.2, 0.5, 0.9$)(12).

Following Cramer et al. (12), we calculated prediction interval (PI) coverage for a set of observations $\left( y_{i}, i=1, \ldots, N \right)$ and PI bounds with an uncertainty level $1-\alpha, \left( l_{\alpha,i}, u_{\alpha,i} \right), i=1,\ldots, N$ as $PI coverage= \frac{1}{N}\sum_{i=1}^{N} 1\left( l_{\alpha,i}\leq y_{i}\leq u_{\alpha,i} \right)$.

**Creating and implementing a naïve model.** To compare the performance of EpiNow2, we generated seven-day forecasts for the same eight time points using a Bayesian generalized linear model (GLM). We implemented the GLM on daily mpox case counts using the stan_glm function in the rstanarm R package (version 2.21.3) (13). We selected the negative binomial family (“neg_binomial_2”) for over-dispersed count data. Our predictors included event date (Table S1, second reference date in column five) and an indicator for weekend, to account for weekend effects in reporting. We considered the GLM to be a “naïve” model because it does not account for epidemiological delay distributions like EpiNow2. We left-truncated the time series, only running the model on the most recent three weeks of data.

Like EpiNow2, stan_glm uses MCMC for implementation (in particular, the highly efficient Hamiltonian Monte Carlo algorithm) (13). We used a prior distribution for the regression coefficients of Normal(0, 2.5) (the package default). The prior distribution for the intercept was Normal(0, 5). We used the default number of MCMC chains (four) and number of iterations (2,000 with the first 1,000 samples discarded as warm-up). Model run time was approximately two seconds. All parameters had R-hat values close to 1, suggesting model convergence.

To obtain the model fits, we used the posterior_predict function in rstanarm on the model object. Out-of-sample predictions (i.e., the forecast) were obtained by using the posterior_predict function on a dataframe containing the seven dates after the end of the right-truncated time series and the indicator for weekend.

To obtain the median and the 20%, 50%, and 90% CrIs, we took the corresponding quantiles of the posterior distribution from the posterior_predict output.

**Supplementary tables**

**Table S1. Input parameters and truncation adjustment for nowcasting/forecasting by outbreak phase.** The difference between the reference dates was used to define the reporting delay. The second reference date listed in column five was used to define the time series (ensuring each case had a non-missing date) and perform the truncation adjustment.

| **Forecast date** | **Outbreak phase** | **Serial interval in days (mean, sd, ref)** | **Incubation period in days (mean, sd, ref)** | **Reference dates** | **Reporting delay distribution (mean, sd)*** | **Truncation adjustment** |
| --- | --- | --- | --- | --- | --- | --- |
| Monday, June 13 | Early | 9.8, 8.6 (14) | 8.7, 4.3 (15) | Rash onset date and OPX test confirmation date | 2.0, 0.5 | Remove 3** most recent days from time series (truncate to Friday, June 10) |
| Monday, June 27 | Early | 9.8, 8.6 (14) | 8.7, 4.3 (15) | Rash onset date and OPX test confirmation date | 1.8, 0.5 | Remove 3** most recent days from time series (truncate to Friday, June 24) |
| Tuesday,  July 5 | Exponential growth | 9.8, 8.6 (14) | 8.7, 4.3 (15) | Rash onset date and event date | 1.8, 0.5 | Remove 4** most recent days from time series (truncate to Friday, July 1) |
| Wednesday, July 27 | Peak | 9.8, 8.6 (14) | 8.7, 4.3 (15) | Rash onset date and event date | 1.9, 0.6 | Remove 5** most recent days from time series (truncate to Friday, July 22) |
| Tuesday, September 6 | Decline | 9.8, 8.6 (14) | 8.7, 4.3 (15) | Rash onset date and event date | 1.9, 0.6 | Remove 4** most recent days from time series (truncate to Friday, September 2) |
| Monday, September 19 | Decline | 9.8, 8.6 (14) | 8.7, 4.3 (15) | Rash onset date and event date*** | 2.0, 0.6 | Remove 3** most recent days from time series (truncate to Friday, September 16) |
| Tuesday, October 11 | Decline | 7.0, 4.2 (10) | 7.5, 4.9 (10) | Rash onset date and event date*** | 1.8, 0.6 | Remove 4** most recent days from time series (truncate to Friday, October 7) |
| Monday, December 5 | Decline | 7.0, 4.2 (10) | 7.5, 4.9 (10) | Rash onset date and event date | 2.0, 0.6 | Remove 3** most recent days from time series (truncate to Friday, December 2) |

OPX: orthopoxvirus.

Event date was determined by the following hierarchy: orthopoxvirus test date, date of call to the call center, date the short CRF was created, and the long CRF timestamp. The hierarchy for event date used for the performance evaluation in December was: diagnosis date, orthopoxvirus test date, orthopoxvirus test confirmation date, case investigation start date, orthopoxvirus sample collection date, date of call to CDC call center, the earliest among report date (to public health department, county, or state), date CDC announced case, and the earliest date the case was entered into DCIPHER, in that order.

*We set the max value at 29 days.

**Including the current day as we ran the models at the end of the day.

***During these two dates, this version of event date was in the process of being deprecated, and the earliest date available for a case was starting to be used.

**Supplementary figures**

**Figure S1. Hierarchy of event dates used for nowcasting/forecasting the 2022 U.S. mpox outbreak.** The data sources available for event date included CDC call center and long and short case report form (CRF), while the data sources for event date (implemented in October 2022) additionally included the National Notifiable Disease Surveillance System. DCIPHER: Data Collection and Integration for Public Health Event Response.

Event date (in use from July - September 2022)

Orthopoxvirus test date

Date of call to call center

Date the short CRF was created

Long CRF timestamp

Event date (in use from October 2022 - December 2022)

Diagnosis date

Orthopoxvirus test date

Orthopoxvirus test confirmation date

Case investigation start date

Orthopoxvirus sample collection date

Date of call to CDC call center

Earliest among report date (to public health department, county, or state)

CDC announced case

Earliest date the case was entered into DCIPHER

**Figure S2. Retrospective evaluation of nowcasts/forecasts of mpox cases in the United States.** Black points correspond to the number of cases in the ground truth data, obtained from DCIPHER on March 16, 2023. The 90% and 50% credible intervals are shown in blue for the EpiNow2 forecasts and green for the GLM forecasts.

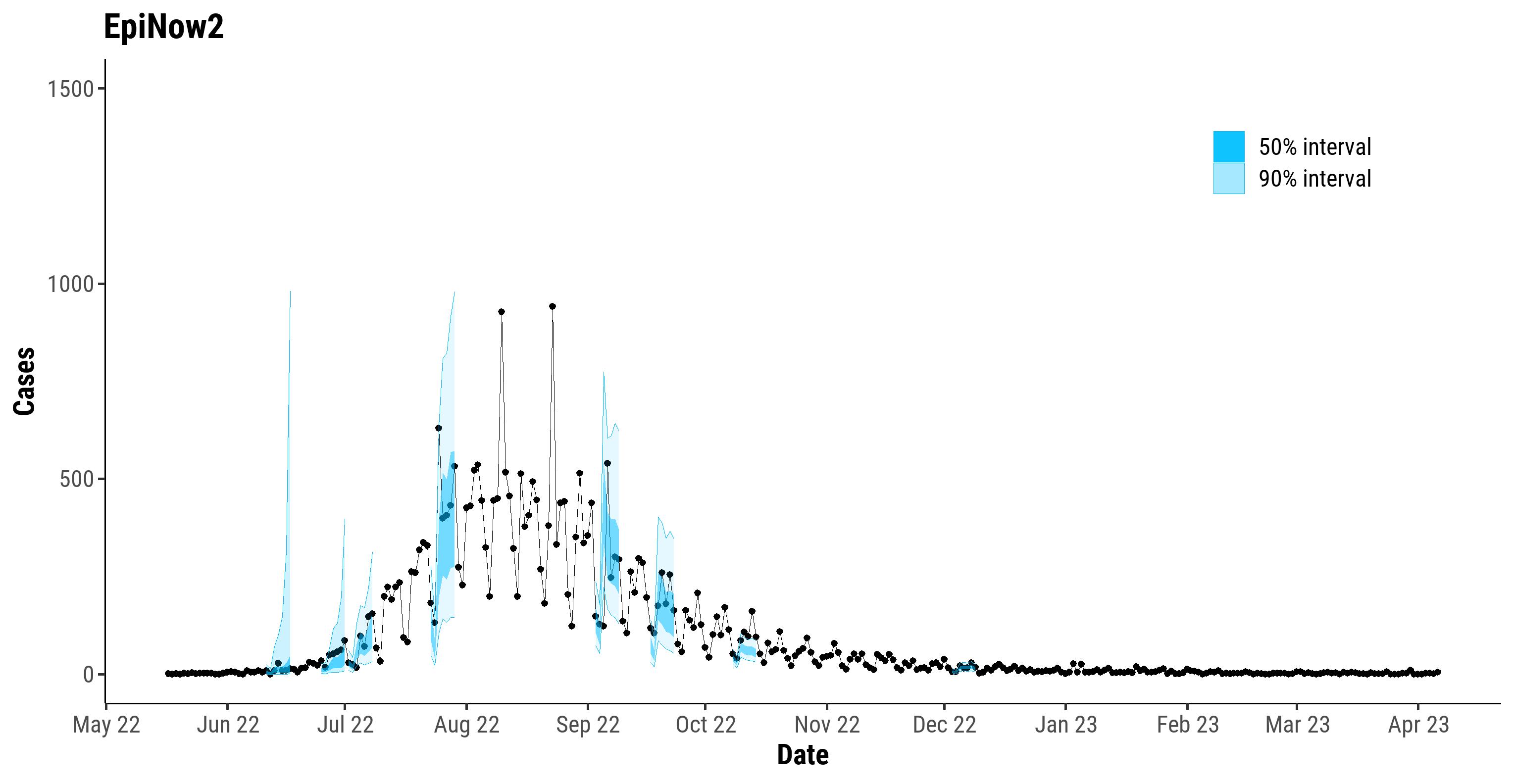

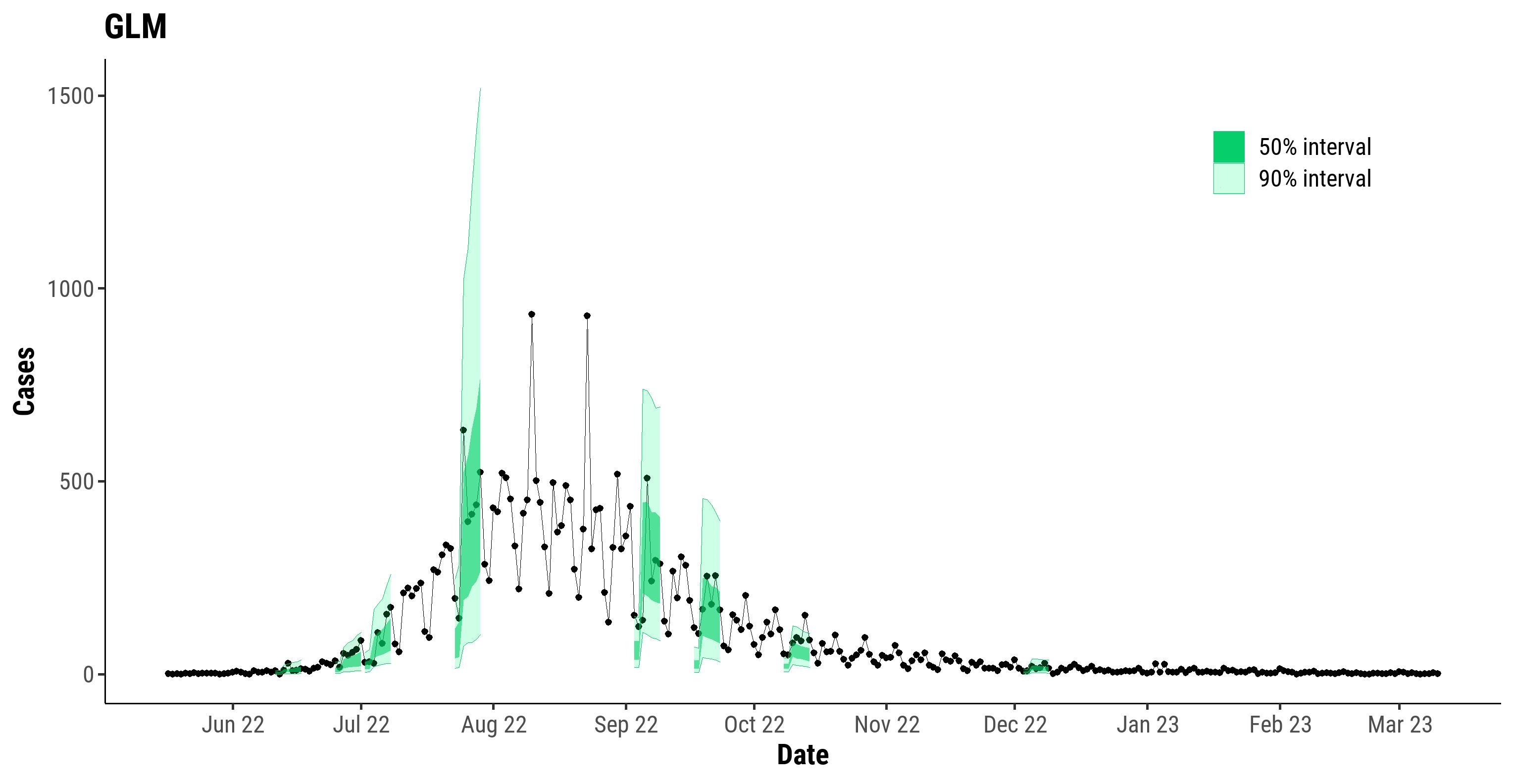

**Figure S3. Time series of reported mpox cases in the United States used for nowcasting/forecasting.** Retrospective evaluation of nowcasts/forecasts were performed for eight dates in the outbreak. Eight of the colored lines in the plot correspond to snapshots of the data available at the time the nowcasts/forecasts were made, while the line labeled “gold” corresponds to the ground truth data, obtained from DCIPHER on March 16, 2023.

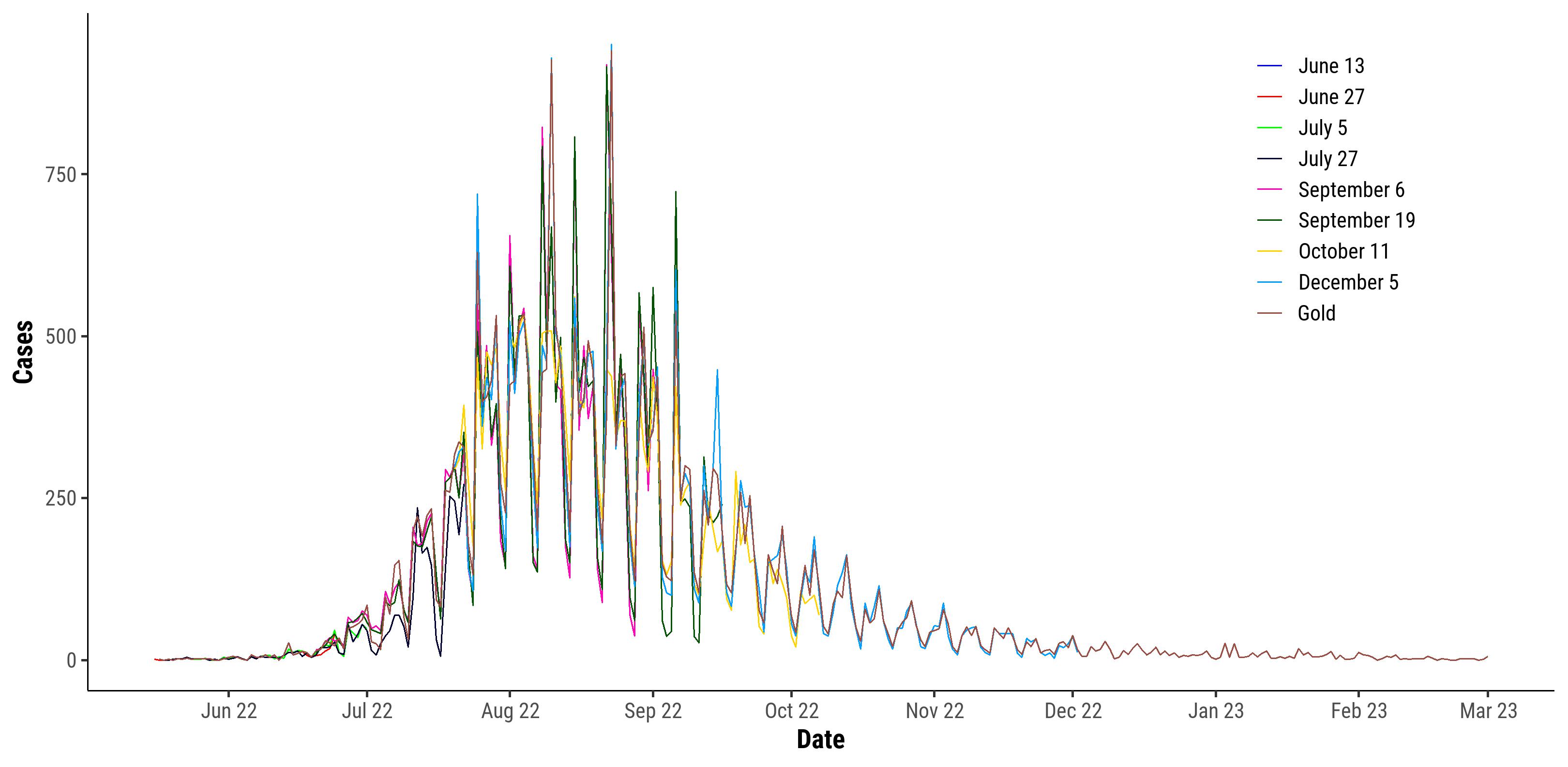
